# The Future of HIV: Challenges in meeting the 2030 *Ending the HIV Epidemic in the U.S. (EHE)* reduction goal

**DOI:** 10.1101/2025.01.06.25320033

**Authors:** Amanda M Bleichrodt, Justin T Okano, Isaac Ch Fung, Gerardo Chowell, Sally Blower

## Abstract

**Objective(s):** To predict the burden of HIV in the United States (US) nationally and by region, transmission type, and race/ethnicity through 2030.

**Methods:** Using publicly available data from the CDC NCHHSTP *AtlasPlus* dashboard, we generated 11-year prospective forecasts of incident HIV diagnoses nationally and by region (South, non-South), race/ethnicity (White, Hispanic/Latino, Black/African American), and transmission type (Injection-Drug Use, Male-to-Male Sexual Contact (MMSC), and Heterosexual Contact (HSC)). We employed weighted (W) and unweighted (UW) *n*-sub-epidemic ensemble models, calibrated using 12 years of historical data (2008-2019), and forecasted trends for 2020-2030. We compared results to identify persistent, concerning trends across models.

**Results:** We projected substantial decreases in incident HIV diagnoses nationally (W: 27.9%, UW: 21.9%), and in the South (W:18.0%, UW: 9.2%) and non-South (W: 21.2%, UW: 19.5%) from 2019 to 2030. However, concerning non-decreasing trends were observed nationally in key sub-populations during this period: Hispanic/Latino persons (W: 1.4%, UW: 2.6%), Hispanic/Latino MMSC (W: 9.0%, UW: 9.9%), people who inject drugs (PWID) (W: 25.6%, UW: 9.2%), and White PWID (W: 3.5%, UW: 44.9%). The rising trends among Hispanic/Latino MMSC and overall PWID were consistent across the South and non-South regions.

**Conclusions:** Although the forecasted national-level decrease in the number of incident HIV diagnoses is encouraging, the US is unlikely to achieve the *Ending the HIV Epidemic in the U.S.* goal of a 90% reduction in HIV incidence by 2030. Additionally, the observed increases among specific subpopulations highlight the importance of a targeted and equitable approach to effectively combat HIV in the US.

## Introduction

In 1981, the Centers for Disease Control and Prevention (CDC) identified the first cases of acquired immunodeficiency syndrome (AIDS) in the United States (US) [1]. However, its cause remained unknown until virologists isolated the first sample of human immunodeficiency virus (HIV) in 1983 [2]. Antiretroviral treatment (ART) regimens have improved considerably throughout the years, enhancing the life-expectancy for those diagnosed with HIV, and subsequently reducing transmission at the population level [3, 4]. Additionally, advances in analytic tools for HIV forecasting have enabled insights into resource demand [5, 6] and future disease burden [7–9] facilitating targeted intervention strategies and policy decisions. Nevertheless, HIV incidence remains alarmingly high and heterogeneous across the US, with 37,981 incident HIV diagnoses reported by the CDC in 2022 [10].

Considerable progress has been made since the formation of the *Ending the HIV Epidemic in the U.S. (EHE)* initiative (2019) in reducing HIV incidence and improving access to care [11]. However, the burden of HIV in the US has remained an inequitable issue across multiple sub-populations [12–14], possibly attributable to continuing racism, HIV-stigma, and disparities in healthcare access [12–16]. Subsequently, the Black/African American community continues to experience a disproportionately high burden of incident diagnoses (38%), followed by Hispanic/Latino (32%) and White (24%) communities [10, 12–15, 17]. Additionally, male-to-male sexual contact (MMSC) is the most common transmission method, followed by heterosexual contact (HSC) and injection drug use (IDU) [10, 17]. While the number of HIV diagnoses among people who reported MMSC or HSC remained stable from 2018 to 2022, HIV diagnoses increased 5% among persons who inject drugs (PWID) during the same period [10, 12, 17]. Regional differences are also noted, with the highest rate of HIV diagnoses in 2022 occurring in the Southern US (15.4%) [10, 13, 15, 17].

Despite the complexity of HIV dynamics being well-characterized in the literature, recent forecasting efforts (since 2016) have lacked sufficient granularity [19–24]. Most studies exploring the future of HIV in the US using real-world data focused on select demographics [18, 22, 23] or sub-populations such as the MMSC [21, 23] and Black/African American communities [21]. Additionally, no identified studies explored the future of HIV at the regional level in the US; however, multiple focused on city-level forecasts [20, 25–27]. Finally, of the identified studies, forecasting horizons varied from one year to 32 years, ranging from 2013 through 2045, and only one study evaluated their forecasts by direct comparison to observed data [19].

An updated and comprehensive overview of the future trajectory of the HIV epidemic in the US is critical to ensure progression towards the *EHE* goal of a 90% reduction in HIV incidence from 2019 to 2030 [11]. Real-time epidemic forecasting can assess the effectiveness of current interventions and policies, thereby providing a bellwether for whether changes in strategy or allocation of resources are needed. When focused on select sub-populations or spatial scales, as HIV-related forecasting often is, concerning trends among non-included groups or locations may be missed, and opportunities for needed intervention and policy tailoring can be overlooked. Thus, timely high-resolution forecasts of the HIV epidemic in the US are essential to capture disease heterogeneity and guide evidence-based policy decisions.

In this report, we conduct multiple forecasts through 2030 of incident HIV diagnoses in the contiguous, Southern, and Non-Southern US, overall and by various races/ethnicities and transmission types. We employ the competitive ensemble *n*-sub-epidemic framework [28] to produce all forecasts in this study which, has shown past success in forecasting COVID-19 [29] and the 2022-2023 mpox epidemic [30, 31], which was also driven by sexual contact [32].

## Methods

### Data Collection and Preparation

We accessed publicly available yearly incident surveillance data regarding the number of HIV diagnoses (2008-2024) and the HIV care cascade in the US from the CDC’s publicly available National Center for HIV, Viral Hepatitis, STD, and TB Prevention *AtlasPlus* dashboard on December 1st, 2024. This database compiles, de-identifies, and aggregates state/local surveillance system data reported to the CDC by demographic and transmission category [33]. Therefore, we could obtain data for the entire US, South, and Non-South regions by transmission category (MMSC, IDU, HSC) and by race/ethnicity (Black/African American, Hispanic/Latino, White). Text S1 provides a detailed breakdown of geographical regions by state (Supplemental Digital Content 1), and the CDC provides further information on data collection details and descriptions of each risk-group in their *Technical Notes* [33].

### Model Calibration and Forecasting

While the number of incident HIV diagnoses is available through 2024, the CDC cautions using data from 2020 and 2021 due to the COVID-19 pandemic, and 2023 and 2024 data remain preliminary [34]. To ensure a consistent time series, we utilized the number of incident HIV diagnoses from 2008 through 2019 for model calibration. To assess the robustness of the 12-year calibration period, we include additional forecasts in Supplemental Digital Content 2 produced using a 14-year calibration period (2008 – 2021) for comparison. Finally, we produced yearly forecasts across transmission and race/ethnicity classifications from 2020 through 2030 nationally, and for the Southern, and non-Southern US.

### The *n*-sub-epidemic framework

We utilized two models, a weighted (W) and unweighted (UW) ensemble, derived from the *n*-sub-epidemic framework [28] to produce all forecasts. The framework aggregates overlapping and asynchronous sub-epidemics to model complex epidemic trajectories. As applied in this analysis, individual sub-epidemics follow the competitive three-parameter generalized logistic growth model and assume a maximum of two-sub-epidemics in the *n*-sub-epidemic trajectory [28]. We used the nonlinear least squares method for parameter estimation and assumed a normally distributed error structure. To quantify parameter and forecast uncertainty, we generated 300 bootstrap samples using the approach described by Hastie et al. [35].

Using the Akaike’s Information Criterion (*AIC_C_*) [28], we selected the two best ranked models based on one or two sub-epidemics. We then derived two ensemble model variations (W & UW) from the top two ranked models to provide different approaches to forecasting HIV’s trajectory. For the weighted model, we used relative likelihood to determine the weights of the top-ranked models and assumed the same weight for both top-ranking models in the unweighted ensemble model [28]. Additional details can be found in Text S2 (Supplemental Digital Content 1) and Chowell et al [28].

All analyses were performed using MATLAB (*R2023a*) software. Examining Model Trends

### Quantifying forecast trends

To quantify the trends noted in each forecast, we used a simple percentage change formula to calculate the relative change (%) in the number of incident HIV diagnoses from 2019 to 2030. The formula is as follows:

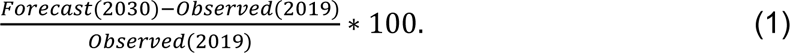

*Forecast (2030)* refers to the forecasted number of incident HIV diagnoses in 2030 and *Observed (2019)* refers to the last year of observed incident HIV diagnoses (2019).

#### Model Uncertainty

We examined the trajectories (i.e., increasing or decreasing) of the upper and lower 95% prediction intervals (95% PI) to provide a qualitative measure of uncertainty.

Additionally, we calculated the average width of the 95% PI across each forecasting period. To calculate the average width of the 95% PI, we first determined the width of the interval for each year included in the forecasting period. We then took the average of the calculated interval widths to determine the average 95% PI width for a given forecasting period.

Finally, we compared results across the weighted and unweighted ensemble models to identify persistent, concerning trends in projected incident HIV diagnosis from 2019 through 2030.

## Results

Figure S1 (Supplemental Digital Content 1) illustrates the uncertainty associated with the incident HIV-diagnosis forecasts nationally and regionally (South and non-South).

**Fig 1.**
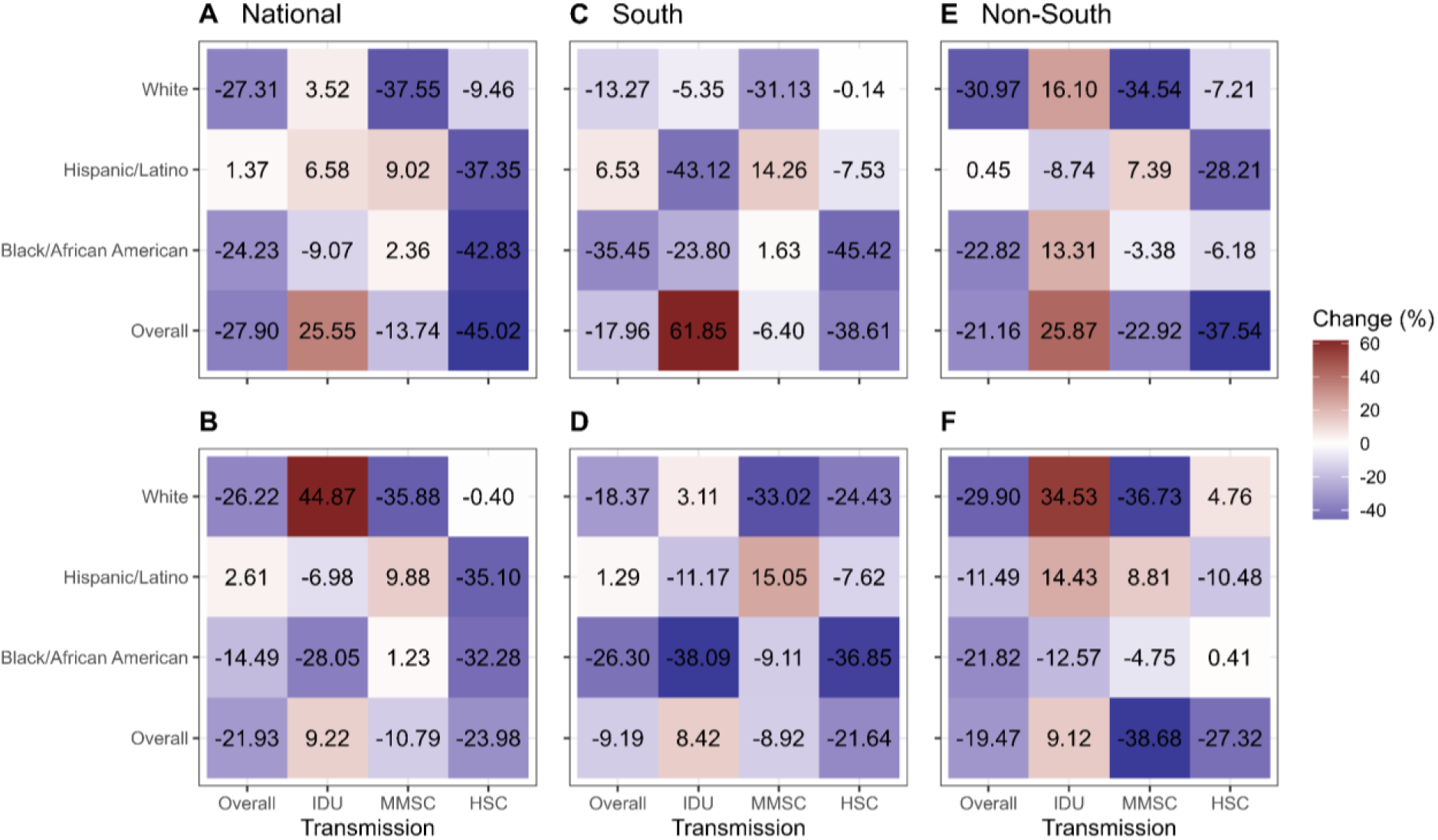
Percent change in the number of incident HIV diagnoses from 2019 to 2030 nationally (A-B), and for the Southern (C-D), and Non-Southern (E-F) United States (US) by transmission category and race/ethnicity. The panel shows the relative percent change from the last year of data used during the model calibration process (2019) to the forecasted median number of incident HIV diagnoses in 2030 nationally (A-B) and for the Southern (C-D), and Non-Southern (E-F) US. Data are provided overall and by race/ethnicity and transmission type: (1) injection drug use (IDU), (2) male-to-male sexual contact (MMSC), (3) heterosexual contact (HSC). Figures A, C, and E show the results from the *n*-sub-epidemic weighted ensemble model, and figures B, D, and F show the results from the *n*-sub-epidemic unweighted ensemble model. Blue tiles indicate forecasted decreases from 2019 to 2030, and red tiles indicate forecasted increases from 2019 to 2030. Darker colors indicate a greater percent change in either direction (i.e., negative or positive). The values within each tile correspond to the relative percent change in incident HIV diagnoses from 2019 to 2030.

### National

#### Overall

Both models forecasted a decrease in the number of incident HIV diagnoses from 2019 to 2030 (W: 27.9% & UW: 21.9%), and among the White (W: 27.3% & UW: 26.2%) and Black/African American (W: 24.2% & UW: 14.5%) communities (Figs. 1-2; Table 1) with moderate levels of precision (Figs. 2, S1). Additionally, both models forecasted stable trends among the Hispanic/Latino community (W: 1.4% & UW: 2.6%) from 2019 to 2030 (Figs. 1-2; Table 1). However, forecast uncertainty was observed for the Hispanic/Latino community, as indicated by fanning in the 95% PIs through 2030 (i.e., increased levels of forecasting uncertainty through 2030) (Figs. 2, S1).

**Fig 2.**
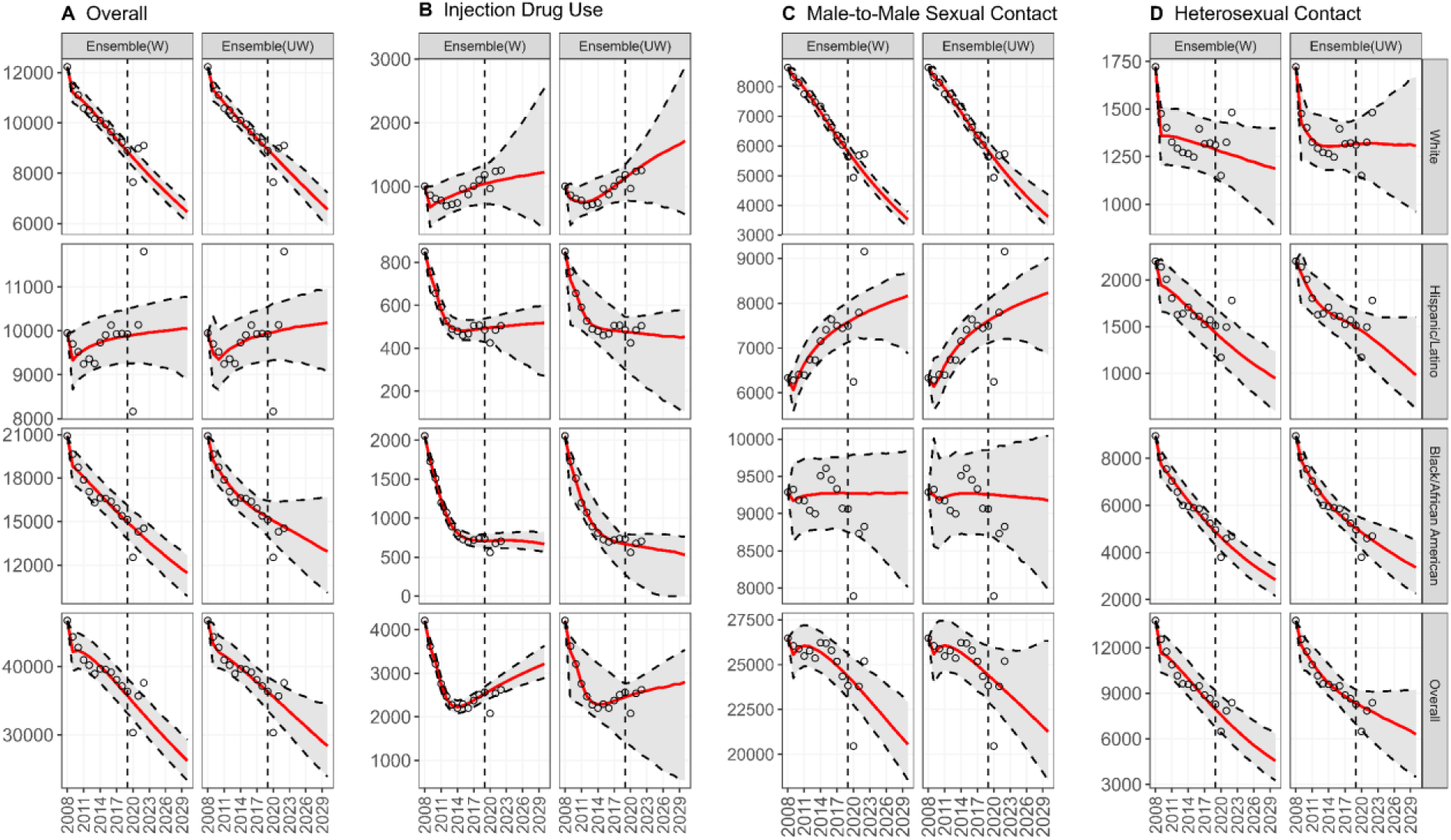
Forecasts for the number of incident HIV diagnoses in the United States (US). The panel shows the forecasts produced using the *n*-sub-epidemic framework (weighted (W) and unweighted (UW) ensemble models) nationally in the US by race/ethnicity and transmission type: (A) overall, (B) injection drug use (IDU), (C) male-to-male sexual contact (MMSC), and (D) heterosexual contact (HSC). The red line is the median forecast, the open circles are the observed data, and the gray ribbon indicates the 95% PI. The vertical dashed line separates the calibration period (left) and forecast period (right).

**Table 1.**
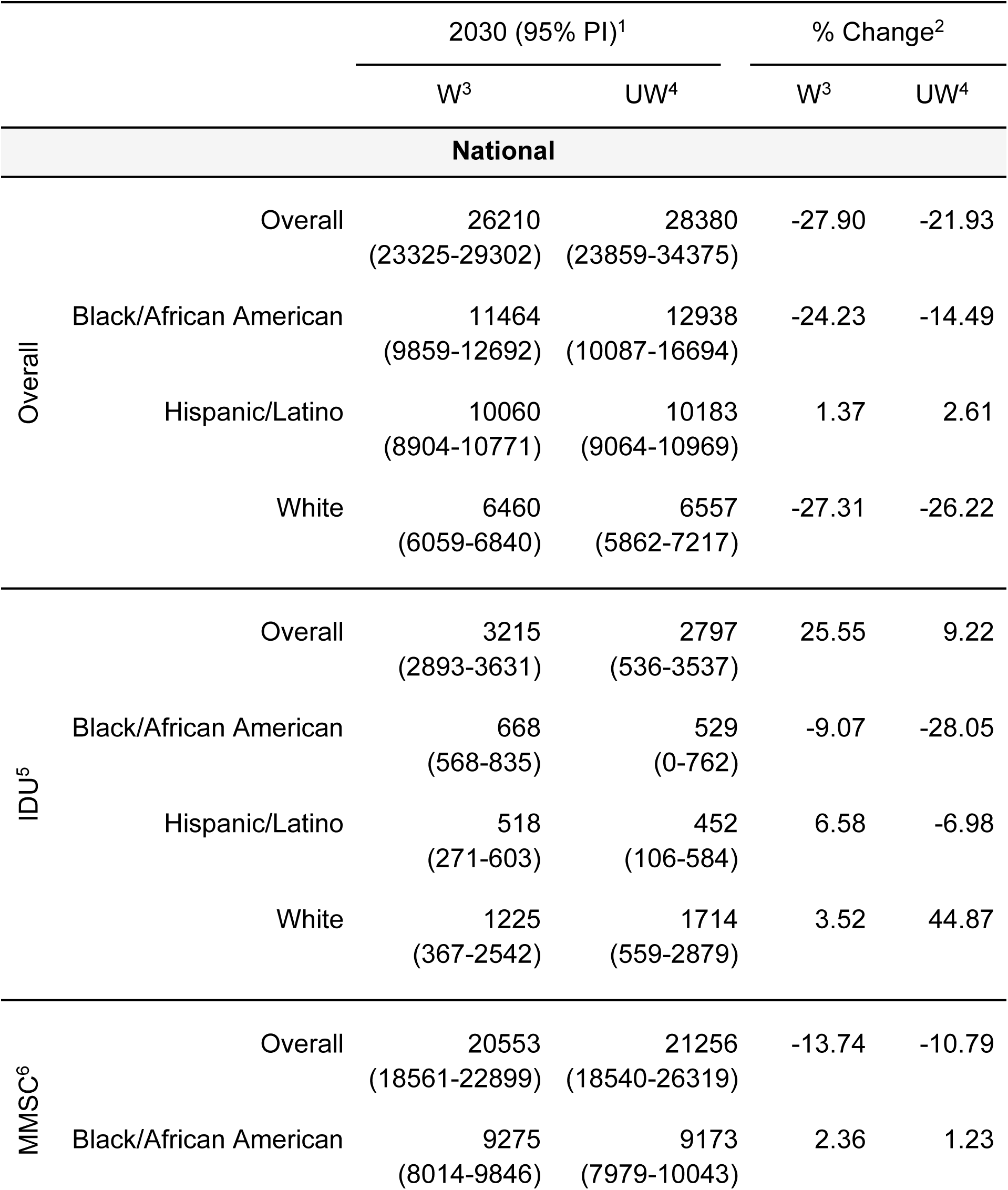
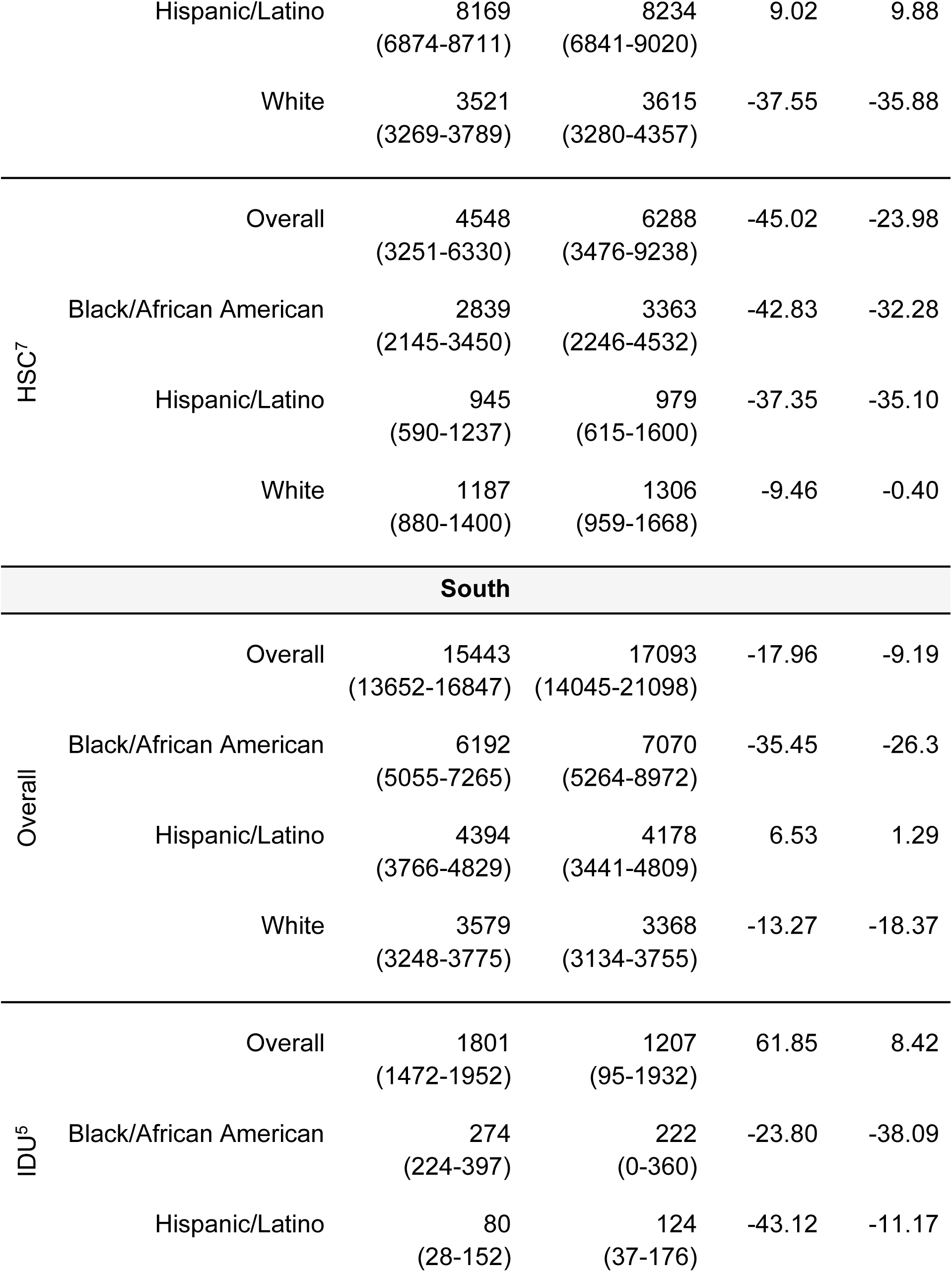
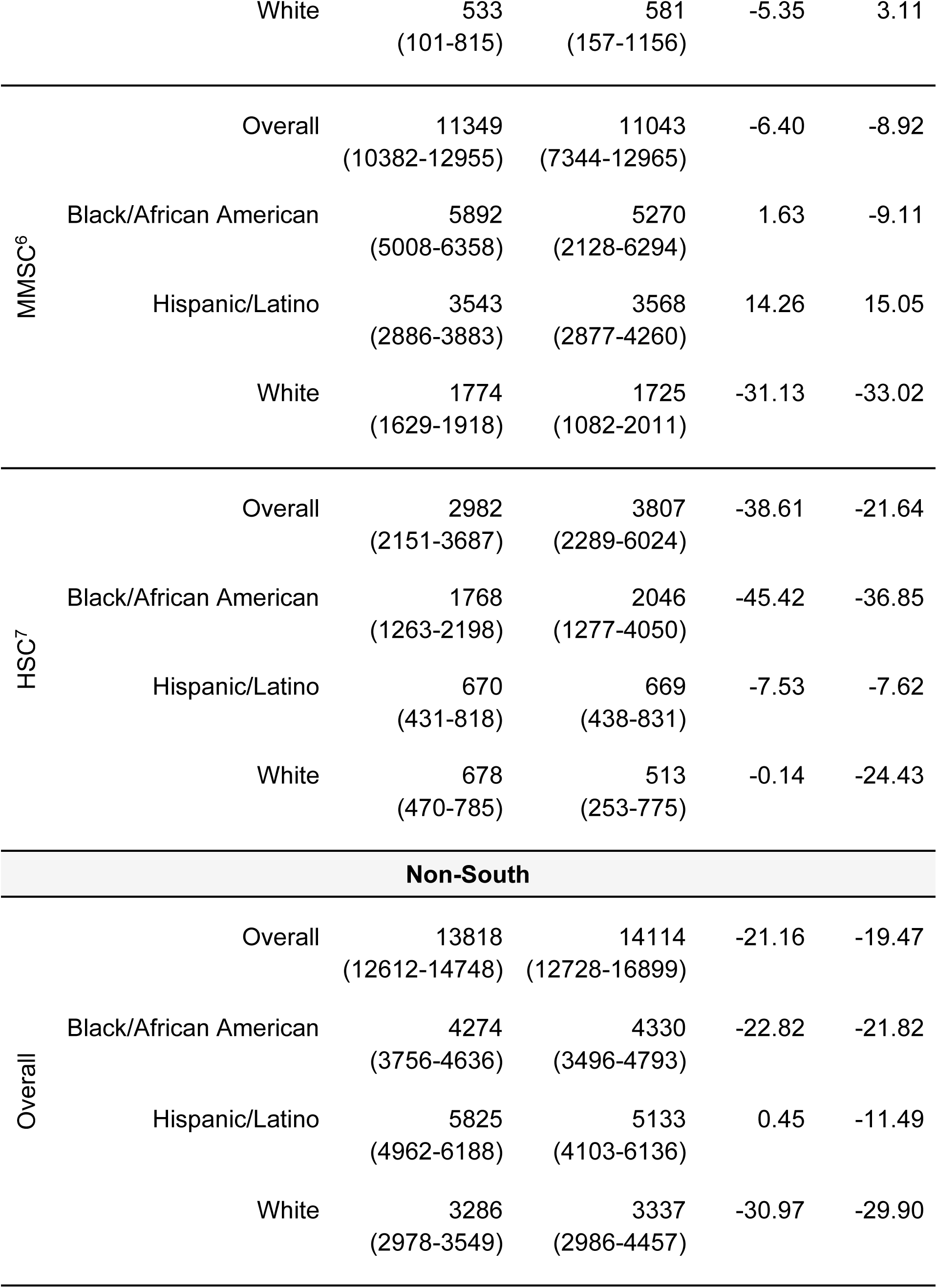
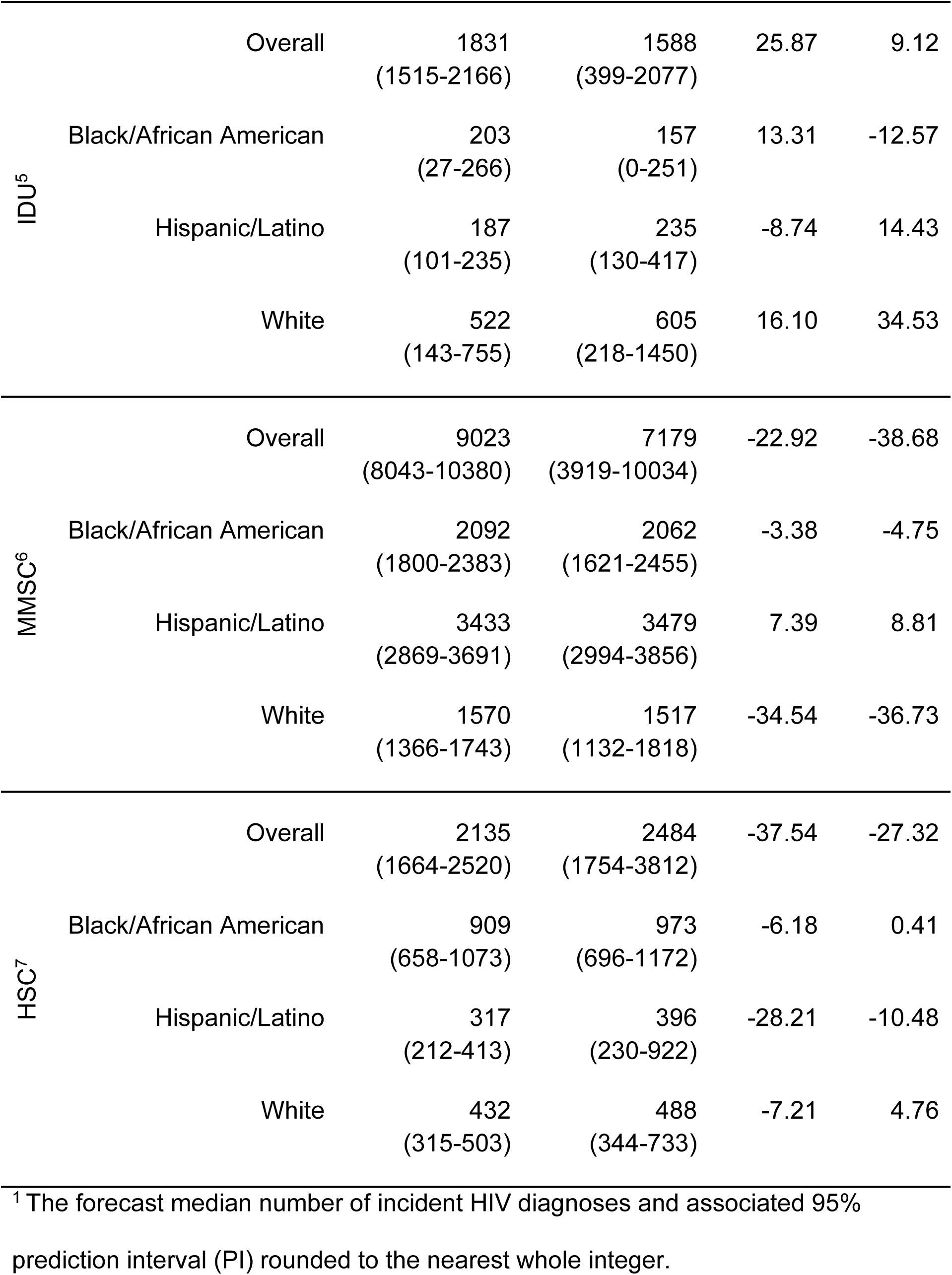
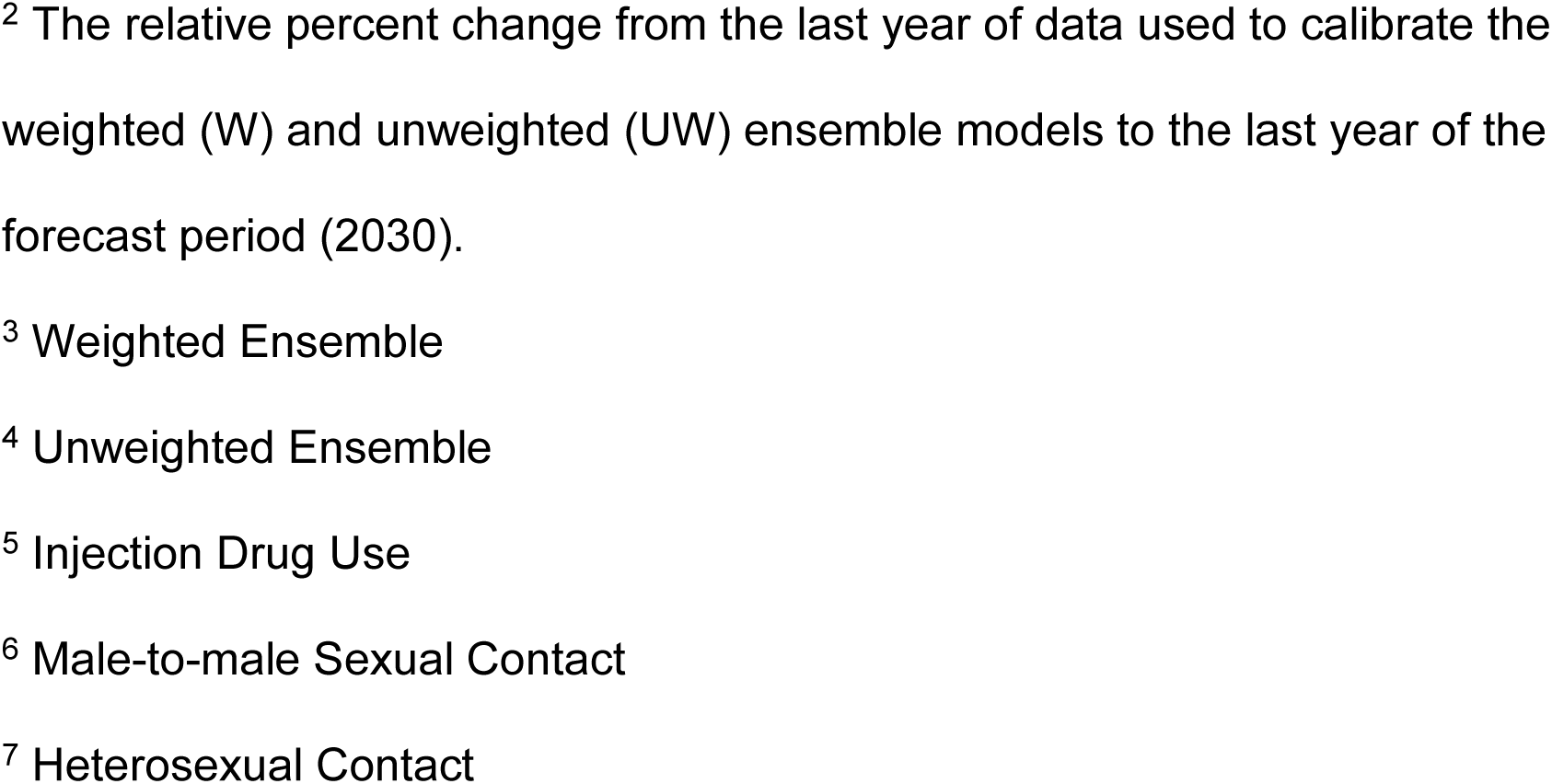
The last year of observed (2019) and forecasted (2030) incident HIV diagnoses and associated percent change from 2019 to 2030 by model, geography, and risk-group.

#### Injection Drug Use (IDU)

Both models predicted increases in the number of incident HIV diagnoses among PWID from 2019 to 2030 overall (W: 25.6% & UW: 9.2%) and among the White community (W: 3.5% & UW: 44.9%) (Figs. 1-2; Table 1). The models forecasted consistent decreases in incident diagnoses among the Black/African American community (W: 9.1% & UW: 28.1%) (Figs. 1-2; Table 1). Nevertheless, most forecasts from both models showed fanning 95% PI bounds (i.e., increased forecasting uncertainty) through 2030, with the weighted ensemble producing more precise forecasts overall (PI Width: 217.4-1355.1) compared to the unweighted ensemble (PI Width: 379.8-2277.8) (Figs. 2, S1).

#### Male-to-Male Sexual Contact (MMSC)

Both models forecasted decreases from 2019 to 2030 in the number of incident HIV diagnoses among people who reported MMSC overall (W: 13.7% & UW: 10.8%) and for the White (W: 37.6% & UW: 35.9%) community (Figs. 1-2; Table 1). The models forecasted consistent increases in the number of incident HIV diagnoses from 2019 to 2030 for the Hispanic/Latino community (W: 9.0% & UW: 9.9%) and stable trends among the Black/African American community (W: 2.4% & UW: 1.2%) (Figs. 1-2; Table 1). However, most forecasts from both models showed more pronounced uncertainty (i.e., fanning), excluding those for the White community (Figs. 2, S1).

#### Heterosexual Contact (HSC)

Both models forecasted decreases in the number of incident HIV diagnoses among people who reported HSC from 2019 to 2030 overall (W: 45.0% & UW: 24.0%) and for all race/ethnicities (Figs. 1-2; Table 1). Nevertheless, the 95% PI trajectories for all weighted ensemble forecasts and only the unweighted ensemble forecasts for the Black/African American and Hispanic/Latino communities followed the decreasing trajectory shown by the median forecast (Figs. 2, S1).

### South

#### Overall

Both models forecasted decreases overall (W: 18.0% & UW: 9.2%) and among the White (W: 13.3% & UW: 18.4%) and Black/African American (W: 35.5% & UW: 26.3%) communities from 2019 to 2030 in the number of incident HIV diagnoses for the South (Figs. 1, 3; Table 1). The models predicted relatively stable trends in incident HIV diagnoses among the Hispanic/Latino (W: 6.5% & UW: 1.3%) community from 2019 to 2030 (Figs. 1, 3; Table 1). The trajectories of the 95% PI bounds for both models followed the median forecast trend only for the White and Black/African American communities (Figs. 3, S1).

**Fig 3.**
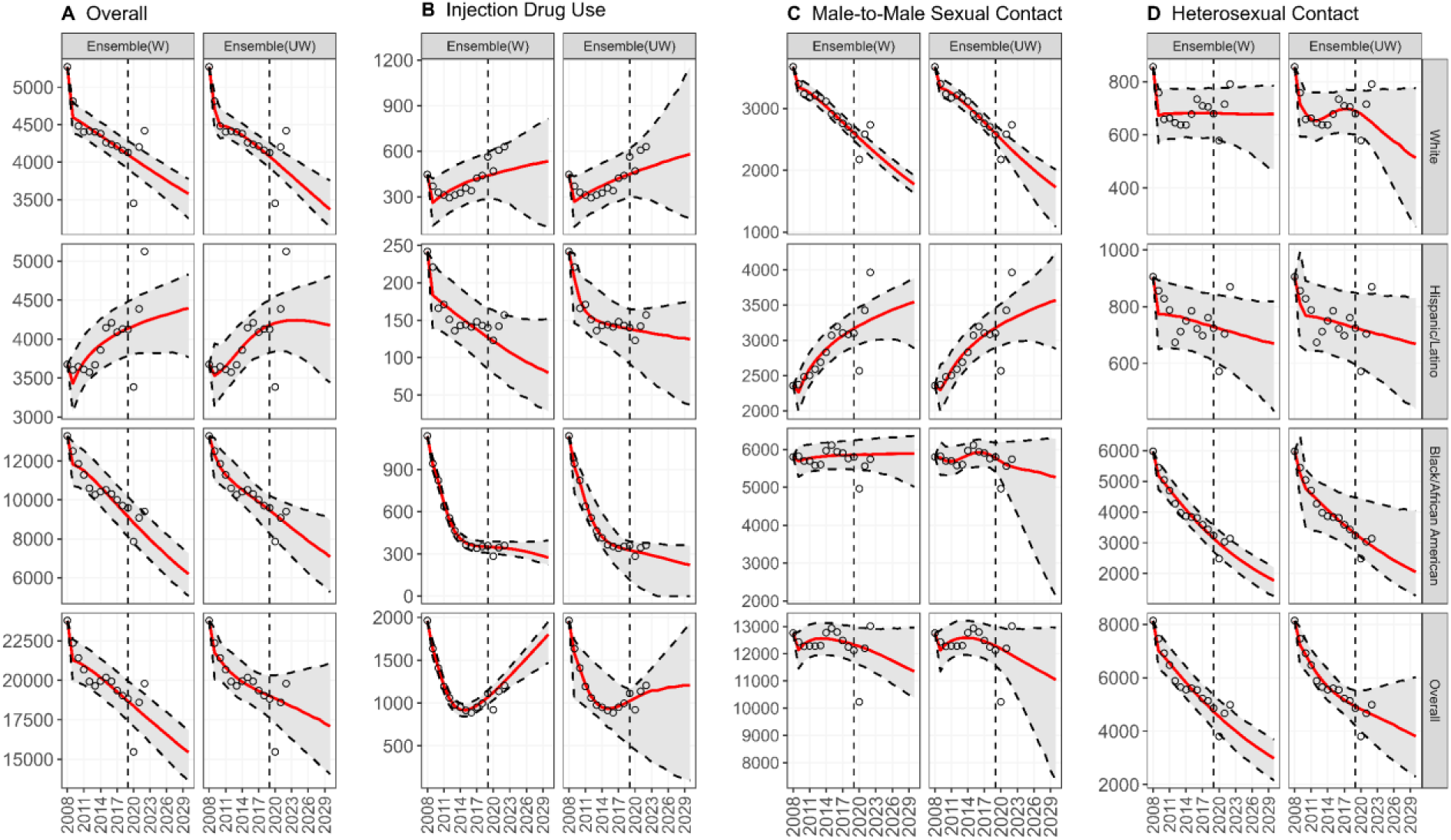
Forecasts for the number of incident HIV diagnoses in the Southern United States (US). The panel shows the forecasts produced using the weighted (W) and unweighted (UW) models derived from the *n*-sub-epidemic framework for the Southern US by race/ethnicity and transmission type: (A) overall, (B) injection drug use (IDU), (C) male-to-male sexual contact (MMSC), and (D) heterosexual contact (HSC). The red line is the median forecast, the open circles are the observed data, and the gray ribbon indicates the 95% PI. The vertical dashed line separates the calibration period (left) and forecast period (right).

#### Injection Drug Use (IDU)

Both models consistently forecasted increases (W: 61.9% & UW: 8.4%) in overall incident HIV diagnoses among PWID, and consistent decreases among the Hispanic/Latino (W: 43.1% & UW: 11.2%) and Black/African American (W: 23.8% & UW: 38.1%) communities from 2019 to 2030 (Figs. 1, 3; Table 1). Most forecasts from both models showed considerable fanning in the 95% PIs through 2030, though the weighted ensemble produced more precise forecasts overall (PI Width: 102.6 – 512.2) compared to the unweighted ensemble model (PI Width: 110.6 – 1306.6) (Fig. 3, S1).

#### Male-to-male sexual contact (MMSC)

Both models forecasted decreases in incident HIV diagnoses among persons who reported MMSC from 2019 through 2030 overall (W: 6.4% & UW: 8.9%) and for the White community (W: 31.1% & UW: 33.0%) (Figs. 1, 3; Table 1). Conversely, the models consistently forecasted increases in the number of incident HIV diagnoses from 2019 to 2030 for the Hispanic/Latino community (W: 14.3% & UW: 15.1%) (Figs. 1, 3; Table 1). Apart from the forecasts for the White community, the remaining forecasts for both models showed considerable fanning in the 95% PIs through 2030 (Figs. 3, S1).

#### Heterosexual Contact (HSC)

Both models forecasted substantial decreases in the number of incident HIV diagnoses among people who reported HSC from 2019 through 2030 overall (W: 38.6% & UW: 21.6%) and for each race/ethnic community (Figs. 1, 3; Table 1). The 95% PI trajectories through 2030 for both models matched the corresponding decrease in median forecasts, except for the overall and White community forecasts (Figs. 3, S1). Additionally, the weighted ensemble produced more precise forecasts overall (95% PI Width: 251.2 - 1416.3) compared to the unweighted ensemble model (95% PI Width: 325.4 – 2668.0) (Fig. S1).

### Non-South

#### Overall

Both models consistently forecasted decreases in the number of incident HIV diagnoses overall (W: 21.2% & UW: 19.5%) and among the White (W: 31.0% & UW: 30.0%) and Black/African American (W: 22.8% & UW: 21.8%) communities in the non-Southern US from 2019 to 2030 with considerable precision (Figs. 1, 4, S1; Table 1). Among the Hispanic/Latino community, the weighted ensemble model forecasted a stable trend (0.5%) in the number of incident HIV diagnoses from 2019 to 2030, whereas the unweighted ensemble model forecasted a decrease (11.5%) during the same period (Figs. 1, 4; Table 1). However, the width of the 95% PI for the weighted ensemble Hispanic/Latino forecasts (PI Width: 927.8) was tighter than that of the unweighted ensemble model (PI Width: 1280.0) (Figs. 4, S1).

**Fig 4.**
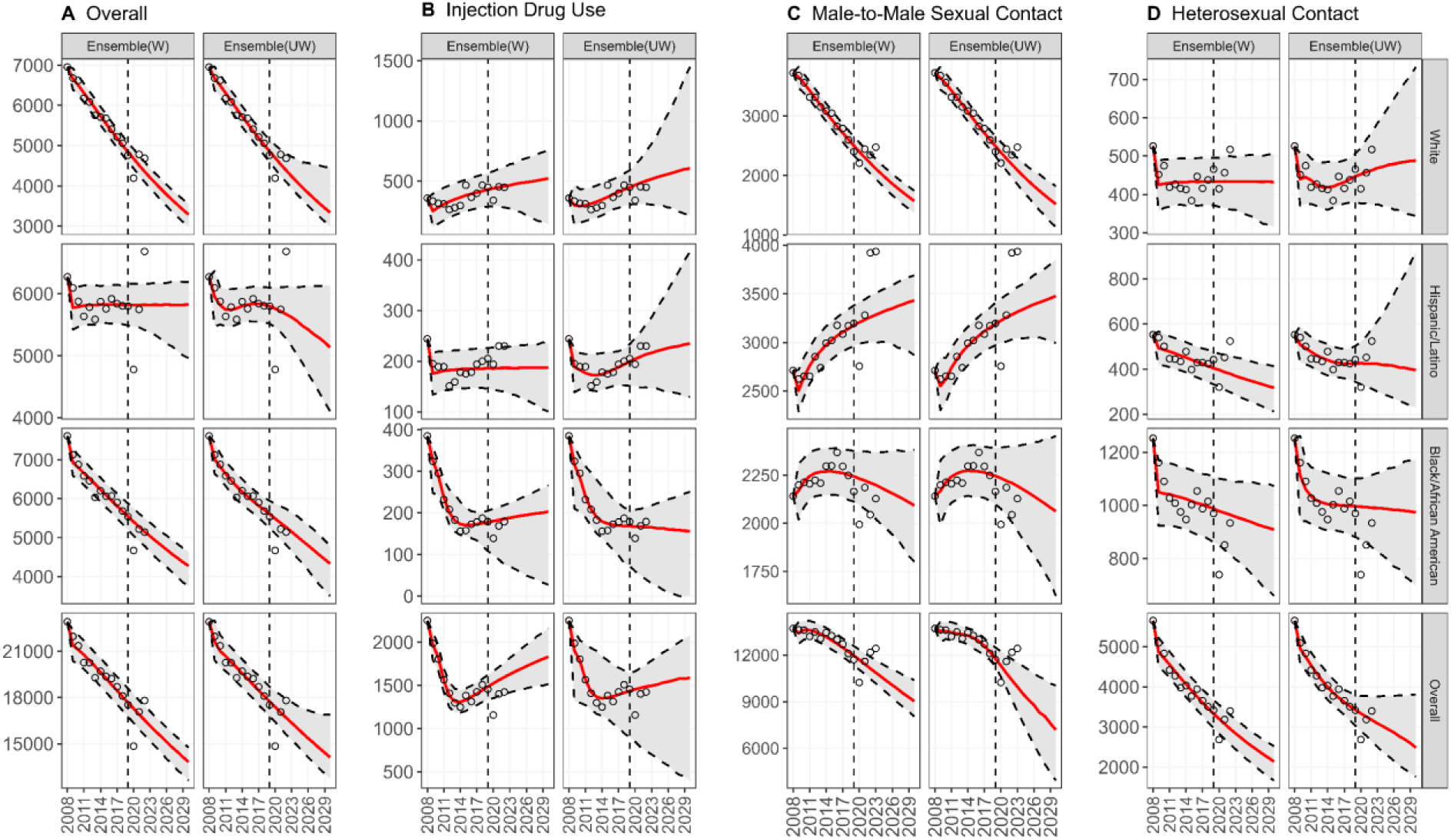
Forecasts for the number of HIV diagnoses in the non-Southern United States (US). The panel shows the forecasts produced using the *n*-sub-epidemic framework (weighted and unweighted ensemble models) for the non-Southern US by race/ethnicity and transmission type: (A) overall, (B) injection drug use (IDU), (C) male-to-male sexual contact (MMSC), and (D) heterosexual contact (HSC). The red line is the median forecast, the open circles are the observed data, and the gray ribbon indicates the 95% PI. The vertical dashed line separates the calibration period (left) and forecast period (right).

#### Injection Drug Use (IDU)

Both models predicted increases in the number of incident HIV diagnoses among PWID from 2019 through 2030 overall (W: 25.9% & UW: 9.1%) and for the White community (W: 16.1% & UW: 34.5%) (Figs. 1, 4; Table 1). However, forecasts conflicted for the Hispanic/Latino and Black/African American communities, and most forecasts from both models showed increased forecasting uncertainty as the trajectory of their 95% PI bounds did not match the corresponding median prediction (Figs. 1, 4, S1; Table 1).

#### Male-to-male sexual contact (MMSC)

Both models forecasted decreases in incident HIV diagnoses among person who reported MMSC from 2019 through 2030 overall (W: 22.9% & UW: 38.7%) and among the White community (W: 34.5% & UW: 36.7%) and stable trends among the Black/African American community (W: 3.4% & UW: 4.8%) (Figs. 1, 4; Table 1). The models forecasted increases among the Hispanic/Latino community (W: 7.39% & UW: 8.81%) (Figs. 1, 4; Table 1). However, both forecasts for the Hispanic/Latino and Black/African American communities showed fanning 95% PI bounds through 2030, and the weighted ensemble model produced a more precise forecasts overall (PI Width: 348.8 – 1750.3) compared to the unweighted ensemble model (PI Width: 519.2 – 4013.5) (Figs. 4, S1).

#### Heterosexual Contact (HSC)

Both models forecasted notable decreases in the number of HIV diagnoses among people who reported HSC overall (W: 37.5% & UW: 27.3%), and for the Hispanic/Latino community (W: 28.2% & UW: 10.5%) (Figs. 1, 4; Table 1). However, for the White and Black/African American communities, the weighted ensemble forecasted small decreases (<10%) in diagnoses from 2019 through 2030, whereas the unweighted ensemble forecasted relatively stable trends (Figs. 1, 4; Table 1). Only the overall forecasts, and those for the Hispanic/Latino and Black/African American communities from the weighted ensemble model produced 95% PIs that consistently followed the trajectory of the median prediction (Figs. 4, S1).

### Additional Results

We provide additional forecasts utilizing the same methods as above but with an extended training period (2008 – 2021) in Supplemental Digital Content 2. We also provide the forecasted trajectory of care cascade uptake through 2030 stratified by sub-population obtained using the 3-parameter GLM [28] in figures S2-S4 (Supplemental Digital Content 1). However, limited data on care cascade elements means these estimates should be interpreted with caution.

## Discussion

Overall, we forecasted a decline from 2019 through 2030 in the overall number of incident HIV diagnoses with considerable certainty for the entire US, South, and Non-South, aligning with findings from previous forecasting efforts [20, 22, 24]. The overall predicted declines are encouraging and highlight the effectiveness of ongoing interventions [36]. However, we are not on track to reach the EHE goal of 90% reduction in HIV incidence by 2030. Additionally, consistent with previous reports [13, 20–22, 24], we forecasted non-decreasing trends in HIV diagnoses through 2030, albeit with some uncertainty, in the following subpopulations: Hispanic/Latino persons overall and those who reported MMSC, White PWID and PWID overall, and nationally among Black/African American persons who reported MMSC. We projected the largest increase in incident HIV diagnoses through 2030 among overall and White PWID. Like Sullivan et al., we also noted regional heterogeneity, with the South forecasted to have the smallest decrease in incident HIV diagnoses compared to the non-South through 2030 [13].

The heterogeneous burden of HIV in the US, identified by this and other studies [13, 20–22, 24], requires focused attention. For example, the forecasted increase in HIV diagnoses among PWID overall poses some concern regarding potential barriers to care [37–39] or perhaps reflects the recent surge in illicit injection drug use [40]. Additionally, highly effective interventions, such as PrEP and Treatment as Prevention can help prevent the transmission of HIV [41–43]. However, well-established racial and ethnic barriers to care have limited the accessibility of such critical tools among those with the highest need [13–15, 44–46]. These disparities also span geographical boundaries, with historically lower access to healthcare resources observed in the Southern US compared to the non-South [13]. While inequitable access to HIV interventions likely contributes to observed heterogeneity, underlying systemic factors, including racism and HIV stigma also play a significant role [11, 13].

Both models produced diverging and variable projections, often with much uncertainty for some sub-populations (Table 1). For example, the weighted ensemble projected an increase from 2019 through 2030 among White PWID in the South, whereas the unweighted ensemble projected a decrease. In other cases, both models forecasted similar directional changes but with varying magnitudes, as observed for overall PWID. These uncertainties likely arise from methodological differences in the models’ construction and complicated interpretation (Text S2, Supplemental Digital Content 1).

Recent literature comparing weighted and unweighted ensembles have found minimal differences in their forecasting performance [31, 47–53]. However, weighted ensembles tended to outperform unweighted ensembles for large-scale outbreaks, whereas unweighted ensembles performed superior with noisier training data [31, 47–53]. Therefore, the weighted ensemble may provide more reliable guidance with stable historical trends, as observed for overall PWID in the South. The unweighted ensemble may be best for forecasting noisier trajectories, as observed among White PWID in the South. We caution readers against overinterpreting the magnitude of projected changes, particularly where data sparsity or methodological differences exacerbate model uncertainties. Instead, we recommend focusing on the average yearly number of diagnoses provided in Table 1 to contextualize differences and guide interpretation when forecasts diverge. This approach ensures that decisions based on these projections consider the inherent challenges posed by sparse data in sub-populations such as PWID.

As a semi-mechanistic approach, the *n*-sub-epidemic framework provides insight into the natural, time-dependent processes that shape the observed epidemic trends through aggregated sub-epidemics [28]. The framework, however, does not explicitly model behavioral modifications, interventions, or other epidemic-shaping factors mechanistically, nor is sensitive to long-term episodic risk behaviors (seasonal or event specific). Therefore, the framework may not capture temporary spikes in diagnoses resulting from short-term changes (i.e., testing events), or alterations to epidemic trajectories that exceed the framework’s assumptions resulting from long-term changes (i.e., improvements in healthcare access). Nevertheless, the framework effectively captured the complex patterns (linear and non-linear) in observed incident HIV diagnoses, and by extension the underlying time-dependent heterogeneities, across sub-populations (Figs. 2-4). Furthermore, the framework has demonstrated superior performance compared to simpler growth and statistical models in forecasting in multiple contexts during which substantial behavioral and policy changes occurred and large uncertainty existed within the available data [29–31].

In 2020, undiagnosed rates were particularly high among PWID, heterosexual women, and the Hispanic/Latino community, followed by an 18% rebound in diagnoses from 2020 to 2021, and a more modest 5% increase from 2021 to 2022 [54, 55]. Therefore, training models through 2019 could result in underprediction of incident HIV diagnoses among disproportionately impacted risk groups. However, inclusion of 2020 and 2021 in the model training process resulted in fewer projected increasing trends through 2030, albeit the increasing trends also observed above for White PWID (national, South, and Non-South), and PWID overall and Hispanic/Latino MMSC community in the South.

Additionally, the primary forecasts outperformed the sensitivity analysis forecasts in capturing the observed number of incident HIV diagnoses in the US in 2022 (Supplemental Digital Content 2). Nevertheless, it is important to note that all incident HIV diagnosis data used for model training must be interpreted with caution as it represents a minimum number of individuals diagnosed annually [33].

While the overall decline in forecasted diagnoses through 2030 is encouraging, the US is unlikely to meet the EHE goal of a 90% reduction in HIV from 2019 to 2030 [12, 25, 27, 56, 57]. The persistent non-decreasing trends forecasted among Hispanic/Latino persons, Hispanic/Latino MMSC, PWID, and White PWID, underscores the importance of a comprehensive approach to HIV forecasting, and intervention and policy formation.

State and local leadership must prioritize increasing access to HIV care for populations disproportionately impacted by HIV, including Black/African American and Hispanic/Latino persons, individuals who report MMSC, and PWID [58]. Public health organizations remain committed to reducing incident HIV diagnoses by 90% by 2030, focusing on disparities in HIV burden across sub-populations [11]. However, to further reduce incidence, it is crucial that ongoing interventions and policies address the diverse landscape of HIV in the US, ensuring universal access to prevention and treatment resources [59].

## Author contributions

**A.M.B.:** Conceptualization, Data curation, Formal Analysis, Visualization, Writing – original draft. **J.T.O.:** Conceptualization, Visualization, Writing – review & editing. **I.C.H.F.:** Conceptualization, Visualization, Writing – review & editing. **G.C.:** Conceptualization, Visualization, Writing – review & editing. **S.B.:** Conceptualization, Visualization, Writing – review & editing.

## Data availability statement

All data used to conduct the analysis are publicly available from the Centers for Disease Control and Prevention’s National Center for HIV, Viral Hepatitis, STD, and TB Prevention *AtlasPlus* dashboard [34]. The associated *SubEpiPredict* toolbox used to conduct all forecasts is also publicly available [28]. The produced HIV forecasts will be made publicly available within a GitHub repository upon acceptance and can be made available upon request.

## Supplemental Digital Content

SupplementalDigitalContent1.pdf SupplementalDigitalContent2.pdf

