## SupplementalDigitalContent1 for "The Future of HIV: Challenges in meeting the 2030 *Ending the HIV Epidemic in the U.S. (EHE)* reduction goal"

**Table of Contents**


| Label | Description | Page(s) |
|---|---|---|
| Text S1 | Geographical Breakdown | 3 |
| Text S2 | The ensemble *n*-sub-epidemic framework. | 5 – 7 |
| Figure S1 | The uncertainty associated with incident HIV-diagnosis forecasts. | 8-9 |
| Figure S2 | Forecasts for the proportion of HIV-positive individuals who meet criteria for each element of the HIV care cascade in the United States. | 10-11 |
| Figure S3 | Percent change in the proportion of individuals meeting the criteria for three elements of the HIV cascade of care from 2019 to 2030 for the United States, by transmission category and race/ethnicity. | 12-13 |
| Figure S4 | The uncertainty associated with forecasts for three HIV cascade of care elements (i.e., linkage to care, receipt of care, and viral level) in the United States. | 14 |

**Text S1. Geographical Breakdown**

Within the main analysis, the United States (US) refers to the 50 states, the District of Columbia, and 5 US territories (American Samoa, Guam, the Northern Mariana Islands, Puerto Rico, and the US Virgin Islands) [1]. The Southern region (South) includes 17 states: Alabama, Arkansas, Delaware, the District of Columbia, Florida, Georgia, Kentucky, Louisiana, Maryland, Mississippi, North Carolina, Oklahoma, South Carolina, Tennessee, Texas, Virginia, and West Virginia [1]. The remaining US states and territories fall under the non-southern regions (non-South) classification.

**Text S2. The ensemble *n*-sub-epidemic framework**

The ensemble *n*-sub-epidemic framework aggregates overlapping and asynchronous growth curves (“sub-epidemics”) to capture complex epidemic trajectories, such as plateaus, resurgences, and waves [2]. The framework has continuously outperformed classic statistical and other simpler growth models in forecasting COVID-19 and mpox [2-5], which is also transmitted primarily through intimate contact [6]. Often mechanistic approaches, such as the commonly employed susceptible-infectious (SI), susceptible-exposed-infected-recovered (SEIR) and other compartmental models are employed in infectious disease forecasting. Such models require the specification or estimation of epidemiological-related parameters, such as transmission rates or infectious periods, to effectively capture and forecast the future burden of disease [7, 8]. Given the sparsity of HIV-diagnosis data for some risk-groups of interest, the complexity of such mathematical models is restricted in terms of the number parameters and mechanisms in which it can estimate. While mechanistic approaches can be tailored to specific situations, the ensemble *n*-sub-epidemic framework provides a previously validated robust approach that does not require the specification of epidemiological parameters. Additionally, the framework does not require extensive training data to produce accurate forecasts, as is characteristic of more complex machine learning and deep learning methodologies [9-11]. Therefore, given the limited nature of HIV-related data in the United States, real-time forecasting using popular data-hungry methods is not often an available methodology. Finally, the “ensemble” feature of the framework also allows for the combination of the predictive power of multiple forecasts, which has shown previous success in producing more accurate forecasts compared to stand-alone models in a variety of contexts [3-5, 12-14].

*The n-sub-epidemic trajectory*

The complete *n*-sub-epidemic trajectory is given by [2]:

$$\frac{dC_i(t)}{dt} = C_i{}'(t) = A_i(t) r_i C_i{}^{p_i}(t)(1 - \frac{C_i(t)}{K_{0i}}). \qquad (1)$$

$C_i(t)$ tracks the cumulative number of incident HIV diagnoses at time $t$, and $r_i, p_i$, and $K_{0i}$ characterize the shape of each sub-epidemic $i$. The parameter $r_i$ gives the growth rate per unit of time and remains positive. The final outbreak size for each sub-epidemic is denoted by $K_{0i}$, and the “scaling of growth” parameter is given by $p_i$. For a given sub-epidemic, when $p_i = 0$, Eq. (1) describes a constant number of incident HIV diagnoses over time whereas when $0 < p_i < 1$, Eq. (1) indicates early sub-exponential growth. If $p_i = 1$, Eq. (1) describes early exponential growth patterns. $A_i(t)$ is an indicator variable used in the timing of the underlying sub-epidemics. However, we used a fixed sub-epidemic onset where all sub-epidemics started at time zero (i.e., 2008).

Our exploratory analysis indicated that a maximum of two-underlying sub-epidemics in the *n*-sub-epidemic trajectory was enough to characterize the observed trends in the data. Additionally, the number of parameters estimated as part of the ensemble *n*-sub-epidemic trajectory scales with the number of sub-epidemics ($3n$) [2]. For example, the framework estimates a maximum of six parameters for each forecast when $n = 2$ (2 sub-epidemics). However, if we assume more than two sub-epidemics ($n > 2$) parameter identifiability becomes problematic as the number of parameters estimated

($\geq 9$) approaches the length of the training period [15]. As the forecasted period is an extension of the model fit, misidentified parameters would lead to erroneous forecast results. Therefore, we assume a maximum of two underlying sub-epidemics to provide ample flexibility in the model fitting process to capture complex epidemiological trajectories while minimizing the risk of parameter misidentification.

*Parameter estimation, model selection and bootstrapping*

We utilized nonlinear least-squares and assumed a normal distributed error structure to estimate model parameters [2]. As the time series data we are fitting to includes relatively large counts, the normal distribution is a good approximation for alternative error structures such as the Poisson or negative binomial distributions [5, 16]. Additionally, normality is a common assumption in infectious disease modeling and forecasting studies [3-5, 16-18].

We employed the corrected Akaike's Information Criterion ($AIC_c$) [2] statistic to help identify the least complex model which best fits the data; this provides insight into whether one or two sub-epidemics should be included. We then selected the two best fit, or ranked, models based on one and two sub-epidemics for each combination of race/ethnicity and transmission type for the national and regional (South and Non-South) levels. The $AIC_C$ for the normal distribution is given by

$$AIC_C = n_d \log(SSE) + 2m + \frac{2m(m+1)}{n_d - m - 1} \tag{2}$$

where $n_d$ is the length of the calibration period and $m$ is the number of model parameters [2]. The parameter $SSE = \sum_{j=1}^{n_d} \left(f\left(t_j, \widehat{\Theta}\right) - y_{t_j}\right)^2$ where $\widehat{\Theta}$ is the set of parameters that minimizes the sum of square differences between the observed data $\{y_{t_1}, y_{t_2}, \dots, y_{t_j}\}$ and the model mean $f(t, \Theta)$ [2].

To quantify parameter and forecasting uncertainty, we generated 300 bootstrap samples assuming normality and using the approach presented in Ref [19]. Additional details regarding the applied parameter estimation and bootstrapping methodologies can be found in Chowell et al. [2].

*Ensemble Model Construction*

We generated two ensembles of the top two ranked models, weighted and unweighted. The weighted ensemble assigns different weights to the top-ranking sub-epidemic models based on their relative likelihoods, where the model with the higher relative likelihood is weighted heavier than that with a lower relative likelihood. The unweighted ensemble weighs each model equally in the ensemble formation. Additional details can be found in Chowell et al. [2].

Weighted ensembles can provide potentially more precise projections, as they place more weight on the better fit of the two top-ranked models based on relative likelihoods. Therefore, the resulting forecasts are largely influenced by the best-fit model, rather than the second-ranked model. In contrast, the unweighted model treats the top-ranking models equally, thus capturing the underlying forecasting uncertainties associated with both included models. This approach can result in broader prediction intervals, particularly when the included models produce divergent forecasts. Nevertheless, these methodological differences highlight the sensitivity of the forecasts to the choice of model and the inherent uncertainties in epidemic modeling.

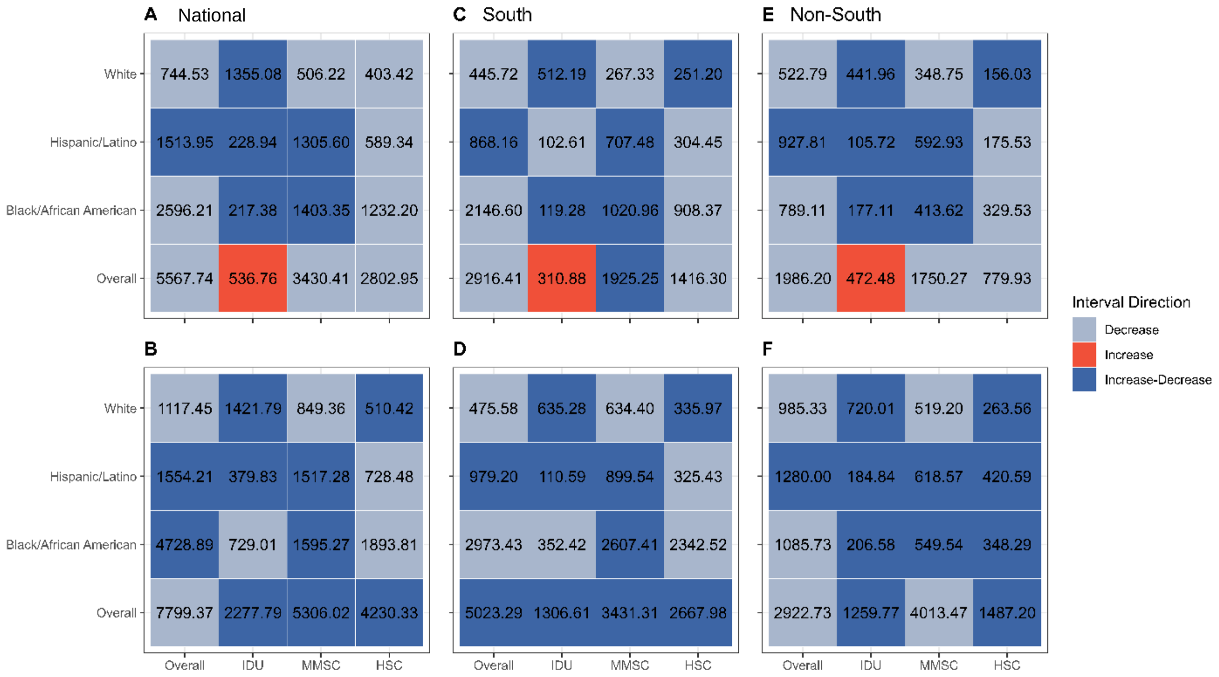


**Fig S1. The uncertainty associated with incident HIV-diagnoses forecasts in the national (A-B), Southern (C-D), and non-Southern (E-F) US.** The panel shows two representations of uncertainty related to incident HIV-diagnosis forecasts (2020 to 2030) generated using the *n*-sub-epidemic framework by transmission category and race/ethnicity for the overall (A-B), Southern (C-D), and non-Southern (E-F) US. The values shown within each tile represent the average 95% PI width across the 11-year forecast period. Data are provided overall and by race/ethnicity and transmission type: (1) injection drug use (IDU), (2) male-to-male sexual contact (MMSC), (3) heterosexual contact (HSC). The top row of figures (A, C, E) shows the uncertainty associated with the *n*-sub-epidemic weighted ensemble model, and the bottom row (B, D, F) shows the uncertainty associated with the *n*-sub-epidemic unweighted ensemble model. The colors shown correspond to the directions of the upper and lower 95% PIs for each forecast. "Decrease" indicates that both bounds decrease from the start to end of the forecasting period, "Increase" indicates both bounds increase from start to end, and "Increase-Decrease" describes contradicting trends between the upper and lower bounds.

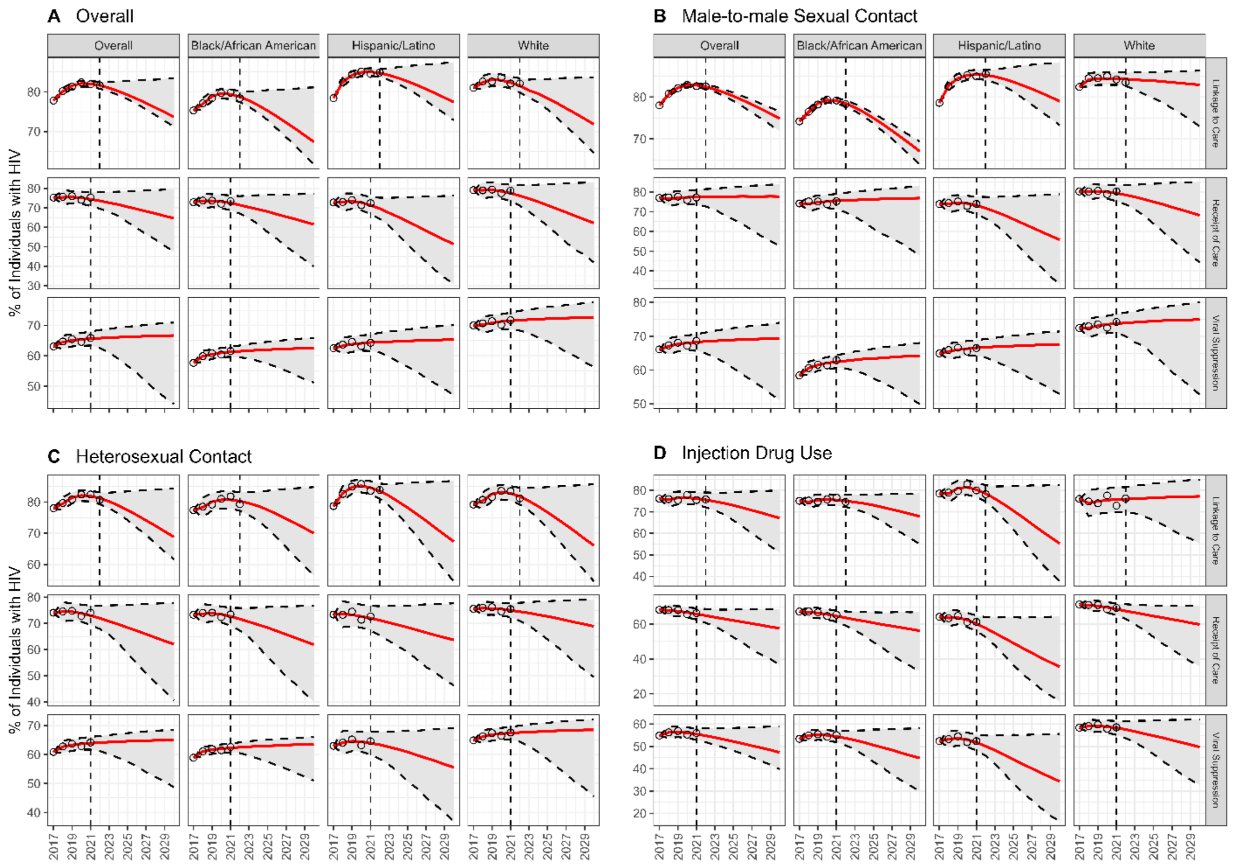


**Fig S2. Forecasts for the proportion of HIV-positive individuals who meet criteria for three elements of the HIV care cascade in the United States.** The panel shows the 8-year (2023-2030) linkage-to-care and 9-year (2022-2030) receipt-of-care and viral suppression forecasts for the percentage of HIV-positive individuals who meet the criteria for each element of the care cascade. Linkage to care is defined as the percentage of individuals diagnosed with HIV who have at least 1 viral load or CD4 test within one month of their diagnosis [20]. Receipt of care is defined as the percentage of individuals diagnosed with HIV who have at least 1 viral load test during 2021 [20]. Viral suppression is defined as the percentage of individuals diagnosed with HIV who have a viral load result of <200 copies/mL at their most recent test [20]. Data are provided by overall (A), race/ethnicity, and transmission type: (B) male-to-male sexual contact

(MMSC), (C) heterosexual contact (HSC), and (D) injection drug use. The linkage-to-care forecasts utilize a 6-year calibration period (2017-2022), and the remaining care-cascade elements use a 5-year calibration period (2017-2021) (i.e., all available data as of December 2024). We conducted forecasts using a generalized logistic model (GLM), assuming normality, for three transmission types, three races/ethnicities, and overall. The red line is the median model fit, and the unfilled circles are the observed data. The grey ribbon is the 95% prediction interval (95% PI) coverage, and the dashed lines that border it correspond to the upper and lower 95% PI. The vertical dashed line is the cut-off between the calibration and forecast periods. Data from 2020 and 2021 needs to be interpreted with caution due to COVID-19. Data was obtained from the Centers for Disease Control and Prevention (CDC) *AtlasPlus* dashboard [21].

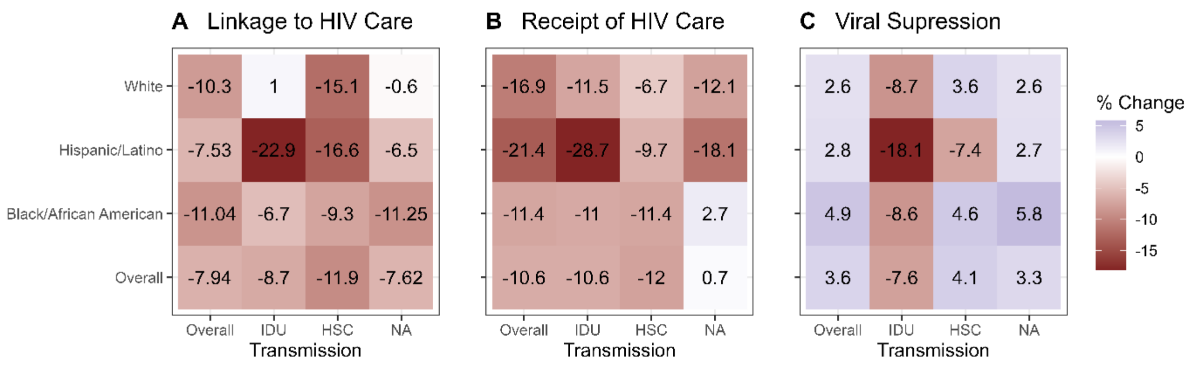


**Fig S3. Percent change in the proportion of individuals meeting the criteria for three elements of the HIV cascade of care from 2019 to 2030 for the United States, by transmission category and race/ethnicity.** The panel shows the absolute percent change in the proportion of individuals from the last year of linkage to care (2022), receipt of HIV care (2021), and HIV viral suppression data (2021) used to calibrate the model, to the forecasted median in 2030 for the United States. The source data corresponds to the percentage of HIV positive individuals who meet the criteria for the given care cascade element (Figure A-C), and is provided by overall, race/ethnicity, and transmission type: (1) injection drug use (IDU), (2) male-to-male sexual contact (MMSC), (3) heterosexual contact (HSC) [1]. Linkage to care is defined as the percentage of individuals diagnosed with HIV who have at least 1 viral load or CD4 test within one month of their diagnosis [20]. Receipt of care is defined as the percentage of individuals diagnosed with HIV who have at least 1 viral load test during 2021 [20]. Viral suppression is defined as the percentage of individuals diagnosed with HIV who have a viral load result of <200 copies/mL at their most recent test [20]. We employed a generalized logistic model (GLM) to produce forecasts and assumed normality. Blue tiles indicate increases from the last observed year of data to 2030, and red indicate decreases during the same time frame. Darker colors indicate larger changes. The values within each tile correspond to the absolute percent change noted overall, and combination of transmission category and race/ethnicity.

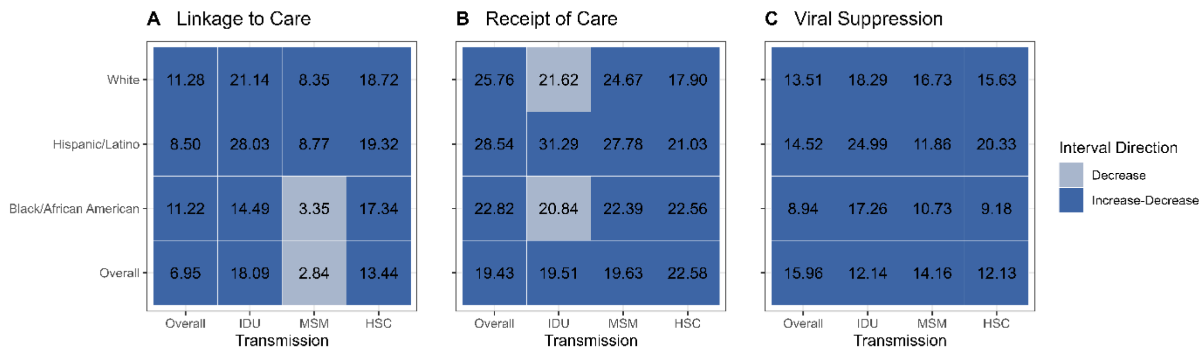


**Fig S4. The uncertainty associated with forecasts for three HIV cascade of care elements (i.e., linkage to care, receipt of care, and viral suppression) in the United States.** The panel shows two representations of uncertainty related to the cascade of care forecasts generated using the generalized logistic model (GLM) by transmission category and race/ethnicity for the entire United States. Linkage to care is defined as the percentage of individuals diagnosed with HIV who have at least 1 viral load or CD4 test within one month of their diagnosis [20]. Receipt of care is defined as the percentage of individuals diagnosed with HIV who have at least 1 viral load test during 2021 [20]. Viral suppression is defined as the percentage of individuals diagnosed with HIV who have a viral load result of <200 copies/mL at their most recent test [20]. Data are provided by overall, race/ethnicity, and transmission type: (1) injection drug use (IDU), (2) male-to-male sexual contact (MMSC), (3) heterosexual contact (HSC). The colors shown correspond to the directions of the upper and lower 95% PIs for each forecast. "Decrease" indicates that both bounds decrease from the start to end of the forecasting horizon, "Increase" indicates both increases from start to end, and "Increase-Decrease" describes contradicting trends between the upper and lower bounds. The values shown within each tile represent the average 95% PI width across the forecast horizon.
