## SupplementalDigitalContent2 for "The Future of HIV: Challenges in meeting the 2030 *Ending the HIV Epidemic in the U.S. (EHE)* reduction goal"

### 1. Purpose

The considerable underdiagnosis of HIV in the United States resulting from the SARS-CoV-2 (COVID-19) pandemic in 2020 and the subsequent rebound in diagnoses in 2021 have been well-established in the existing literature [1-3]. However, as discussed by the Centers for Disease Control and Prevention (CDC), the 5% increase in HIV diagnoses observed from 2021 to 2022 is only slightly higher than the expected 2%-3% change expected each year based upon pre-pandemic data [1]. Given the known effect of COVID-19 on reported diagnoses rates in 2020 and 2021, we elected in our primary analysis to include data only through 2019 in our model calibration process. Nevertheless, we acknowledge that the disruptions in clinical care, patient hesitancy in accessing care, and testing material shortages in 2020 may continue to impact HIV diagnoses for years to come [1].

As data is now available through 2022 (2023 and 2024 data are still considered preliminary), evaluating the short-term performance of our forecasts is critical to ensure that our models are accurately capturing the future burden of disease in a post-COVID-19 society. Additionally, a thorough exploration of the potential effect of COVID-19 impacted data on our projections can provide additional support for stopping our model training period in 2019. Therefore, this brief analysis aims to determine the accuracy of the forecasts produced utilizing the methodology presented in our primary manuscript and Supplemental Digital Content 1. We also compare our primary results, including percent change from 2019 to 2030 and 1-year forecasting performance (2022) to forecasts produced using the same framework but with an extended calibration period (2008 through 2021).

### 2. Methods

We utilized the same methods presented in the main manuscript and Text S2 (Supplemental Digital Content 1) to conduct additional forecasts now trained with data from 2008 through 2021, rather than from 2008 through 2019, as done for the primary forecasts.

*Model Evaluation & Comparison*

To determine the short-term forecasting performance of the forecasts trained with data from 2008 to 2019 and 2008 to 2021, we calculated the mean squared error (MSE), mean absolute error (MAE), and 95% prediction interval coverage (95% PI). We evaluated both methodologies' performance in forecasting incident HIV diagnoses in 2022, the last year for which non-preliminary data was available from the CDC's National Center for HIV, Viral Hepatitis, STD, and TB Prevention *AtlasPlus* dashboard [4]. Additional details regarding the applied statistics can be found in [5, 6].

We also compared the corresponding percent change (2019 to 2030) results between forecasts produced from the ensemble *n*-sub-epidemic framework trained with data through 2019 and those from the framework trained with data through 2021.

### 3. Results

*Forecasting Results*

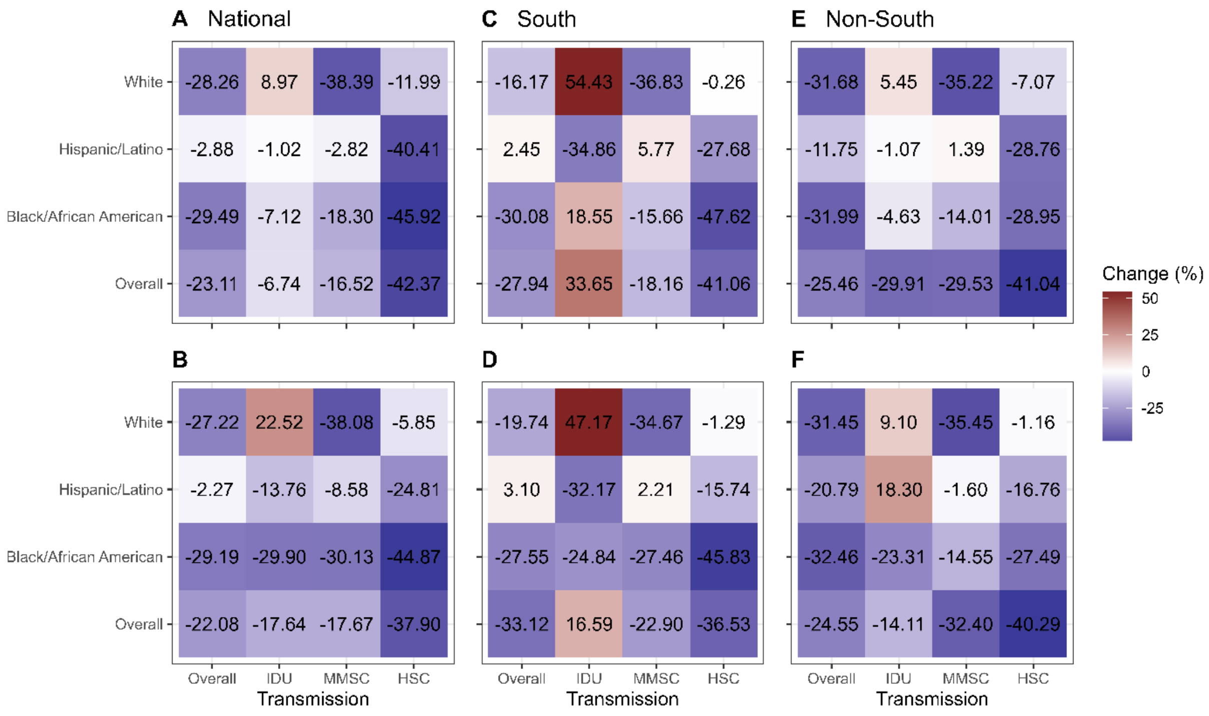


**Fig 1S. Percent change in the number of incident HIV diagnoses from 2019 to 2030 nationally (A-B), and for the Southern (C-D), and Non-Southern (E-F) US by transmission category and race/ethnicity.** The panel shows the relative percent change from the last year of data used during the model calibration process (2021) to the forecasted median number of incident HIV diagnoses in 2030 for the overall (A-B), Southern (C-D), and Non-Southern (E-F) US. Data are provided overall and by race/ethnicity and transmission type: (1) injection drug use (IDU), (2) male-to-male sexual contact (MMSC), (3) heterosexual contact (HSC). Figures A, C, and E show the results from the *n*-sub-epidemic weighted ensemble model, and figures B, D, and F show the results from the *n*-sub-epidemic unweighted ensemble model. Blue tiles indicate forecasted decreases from 2019 to 2030, and red tiles indicate forecasted increases from 2019 to 2030. Darker colors indicate a greater percent change in either direction (i.e., negative or positive). The values within each tile correspond to the relative percent change in incident HIV diagnoses from 2019 to 2030.

Overall, the forecasts produced from the ensemble *n*-sub-epidemic framework trained with data through 2021 predicted consistent decreases for most geographical scales, transmission categories, and race/ethnicities from 2019 to 2030. However, both models (weighted and unweighted) forecasted considerable increases among the White persons who inject drugs (PWID) for all three spatial scales (5.5% to 54.4%), as well as for PWID overall (16.6% to 33.7%), the Hispanic/Latino community overall (2.5% to 3.1%) and who reported male-to-male sexual contact (MMSC) (2.2% to 5.8%), in the Southern United States.

Compared to the forecasts included within the primary analyses, the forecasts produced using the extended training period (2008 – 2021) predicted less frequent increases from 2019 through 2030 across transmission types, race/ethnicities, and geographies. Additionally, when decreases were projected from 2019 to 2030, those presented in Figure 1S were more frequently larger in scale compared to the forecasts produced using training data from 2008 through 2019. A summary of the major differences between the methodology utilized in the main analysis and presented in this sensitivity analysis is included within Table 1S below.

**Table 1S.** A summary of the major differences, including the number of forecasts in which increases and decreases occurred, and magnitude of change from 2019 through 2030, utilizing two different calibration periods.

| **Direction of Projected Trend (2019 – 2030)[1]** | | |
|---|---|---|
| | Main Analysis (2008 – 2019) | Sensitivity Analysis (2008 – 2021) |
| Increase | 30 | 15 |
| Decrease | 66 | 81 |
| **Magnitude of Change (2019 – 2030)[2]** | | |
| | Main Analysis (2008 – 2019) | Sensitivity Analysis (2008 – 2021) |
| Increase | 0.41% – 61.85% | 1.39% - 54.43% |
| Decrease | 0.14% – 45.02% | 0.26% - 45.92% |

[1] The number of forecasts the specified trend (increasing or decreasing) occurred when comparing the observed number of incident HIV diagnoses across geographies, transmission types, and race/ethnicities in 2019 to the forecasted median value in 2030. A total of 96 forecasts were conducted for each calibration period length.

[2] The resulting range in percent changes values from 2019 through 2030 across geographies, transmission types, and race/ethnicities for each observed trend classification.

The forecasts included as part of the sensitivity analyses forecasted decreases among overall PWID on the National level and in the non-southern regions of the United States, whereas the forecasts in the main analysis predicted increases from 2019 to 2030. Additionally, the main analysis identified concerning non-decreasing trends among the Hispanic/Latino community overall and among Hispanic/Latino people who reported MMSC across all three geographical scales. The forecasts presented in Figure 1S projected consistent increases among the same communities only in the Southern United States.

*Model Evaluation*

Compared to the forecasts included within the main analysis, those included within the sensitivity analyses performed worse overall (across performance metrics) in capturing the one-year ahead forecasts (2022). Table 2S below presents the frequency of “success”, or performing best for a given metric, of both the weighted and unweighted ensemble models and analyses employed. Successes were aggregated by location; therefore, each location has 16 associated scenarios (i.e., transmission types and race/ethnicities). The forecasts produced using the unweighted ensemble *n*-sub-epidemic model calibrated with data from 2008 through 2019 performed best most frequently across locations, transmission types, and race/ethnicities.

**Table 2S.** The frequency of “success”[1] in MSE, MAE, and 95% PI for each model, analysis, and geographical region of interest.

| | | | | |
|---|---|---|---|---|
| **Mean Squared Error (MSE)** | | | | |
| | Main Analysis (2008 – 2019) | | Sensitivity Analysis (2008 – 2021) | |
| | Weighted | Unweighted | Weighted | Unweighted |
| National | 12.50% (2/16) | **75.00% (12/16)** | 6.25% (1/16) | 6.25% (1/16) |
| South | **50.00% (8/16)** | 43.75% (7/16) | 6.25% (1/16) | 0.00% (0/16) |
| Non-South | 37.50% (6/16) | **43.75% (7/16)** | 0.00% (0/16) | 18.75% (3/16) |
| **Mean Absolute Error (MAE)** | | | | |
| | Main Analysis (2008 – 2019) | | Sensitivity Analysis (2008 – 2021) | |
| | Weighted | Unweighted | Weighted | Unweighted |
| National | 12.50% (2/16) | **75.00% (12/16)** | 6.25% (1/16) | 6.25% (1/16) |
| South | **50.00% (8/16)** | 43.75% (7/16) | 6.25% (1/16) | 0.00% (0/16) |
| Non-South | 37.50% (6/16) | **43.75% (7/16)** | 0.00% (0/16) | 18.75% (3/16) |
| **Prediction Interval Coverage (95% PI)[2]** | | | | |
| | Main Analysis (2008 – 2019) | | Sensitivity Analysis (2008 – 2021) | |
| | Weighted | Unweighted | Weighted | Unweighted |
| National | 75.00% (12/16) | **93.75% (15/16)** | 81.25% (13/16) | **93.75% (15/16)** |
| South | 62.50% (10/16) | **93.75% (15/16)** | 68.75% (11/16) | 87.5% (14/16) |
| Non-South | 56.25% (9/16) | **100% (16/16)** | 75.00% (12/16) | 87.5% (14/16) |

[1] A “success” is granted when the model produced either the lowest MSE or MAE, or highest 95% PI coverage for a given geography, transmission type, and race/ethnicity.

[2] Row totals for 95% PI coverage may exceed *n = 16* due to ties between models. If a tie (i.e., equally high 95% PI coverage), occurred both models were awarded a success.

### 4. Key Takeaways

The forecasts generated using the ensemble *n*-sub-epidemic framework, trained with data from 2008 to 2021, produced slightly different trends compared to those presented in the primary manuscript. Notably, the widespread non-decreasing trends projected among PWID, the Hispanic/Latino community overall, and Hispanic/Latino individuals reporting MMSC in the primary analysis were not observed in the sensitivity analysis. Furthermore, the magnitude of change for decreasing trends was generally larger in the sensitivity analysis compared to the main analysis. However, the forecasts generated using the ensemble *n*-sub-epidemic framework calibrated with data from 2008 to 2019 outperformed those trained with data through 2021 in capturing the observed number of incident HIV diagnoses in the United States in 2022. Given the literature-backed increases in incident HIV diagnoses among high-risk populations (i.e., PWID, Hispanic/Latino community) [1, 7-11] and the successful short-term forecasting performance of the primary analysis methodology, this sensitivity analysis supports exclusion of 2020 and 2021 during our model training process.
